# Integrative genomics analysis implicates decreased *FGD6* expression underlying risk of intracranial aneurysm rupture

**DOI:** 10.1101/2022.03.12.22272299

**Authors:** Andrew T. Hale, Jing He, Jesse Jones

## Abstract

**Background:** The genetic determinants and mechanisms underlying intracranial aneurysm rupture (rIA) are largely unknown. Given the ∼50% mortality rate of rIA, approaches to identify patients at high-risk will inform screening, diagnostic, and preventative measures.

**Objective:** Our goal was to identify and characterize the genetic basis of rIA.

**Methods:** We perform a genome-wide association study (GWAS) use functional genomics approaches to identify and characterize rIA-associated loci and genes. We perform a meta-analysis across 24 published GWAS of rIA. Single nucleotide polymorphisms (SNP), gene-burden analysis, and functional genomics identify and characterize genetic risk factors for rIA.

**Results:** Our cohort contains 84,353 individuals (7,843 rIA cases and 76,510 controls). We identify 5 independent genetic loci reaching genome-wide significance (p<5.0×10^−8^) for rIA including rs12310399 (*FGD6*, OR=1.16), which to our knowledge, has not been implicated in prior GWAS of rIA. We then quantified gene-level mutation-burden across ∼20,000 genes, and only *FGD6* (containing 21 rIA-associated SNPs) reached transcriptome-wide significance. Expression quantitative trait loci (eQTL) mapping indicates that rs12310399 causes decreased *FGD6* gene expression in arterial tissue. Next, we utilized publicly available single-cell RNA sequencing of normal human cerebrovascular cells obtained during resection surgery and identify high expression of *FGD6* in 1 of 3 arterial lineages but absent in perivascular cells. These data suggest how alterations in FGD6 may confer risk to rIA.

**Conclusion:** We identify and characterize a previously unknown risk loci for rIA containing *FGD6*. Elucidation of high-risk genetic loci may instruct population-genetic screening and clinical-genetic testing strategies to identify patients predisposed to rIA.

**Funding:** No funding sources were used for the material presented herein.

## Introduction

Intracranial aneurysms (IA) are abnormal dilations of cerebral arteries that occur in response to vessel wall dysfunction. Spontaneous rupture of an IA (rIA) causes severe neurological injury and death.^1,2^ IA is estimated to affect around 5% of the world’s population with lifetime rupture-risk of ∼25%.^3^ However, the mortality of rIA is ∼50%.^4^ While many rIA risk-factors including family history, female gender, aneurysm location/size, cigarette smoking, and hypertension have been elucidated in large epidemiological studies,^5^ these factors are largely non-specific. Thus, personalized genetic approaches to 1) identify patients with IA, and 2) predict which patients with IA are most likely to rupture are needed.

Elucidating causal genetic factors associated with rIA has direct implications for disease prevention, diagnosis, and treatment. While rare inherited causes of IA have been described,^6^ these patients comprise the minority of total IA disease burden.^7^ Family history of rIA in first-degree relatives, even in the absence of a known IA-associated genetic syndrome, increases one’s risk of being diagnosed with rIA by nearly four-fold.^8^ Thus, a strong genetic component is thought to underlie IA,^9^ and unbiased, genome-wide approaches to understand the germline-genetic determinants of IA formation and rupture are critical to understanding IA pathophysiology and quantification of rupture risk. Since one’s genetic information in large part does not change over a lifetime (excluding somatic mutations), elucidating the genetic architecture of rIA can inform screening, detection, and treatment (for patients at the highest risk) strategies. In this study, we systematically delineate the genetic determinants of rIA using convergent human-genetic and functional genomics approaches.

## Methods

### Genome wide association study analysis

Genetic data were obtained using the Cerebrovascular Disease Knowledge Portal (cerebrovascularportal.org).^10^ GWAS was performed as previously described.^11,12^ Whole-genome genotyping was performed using Illumina HumanOmniExpressExome BeadChip.^10^ Imputation was performed using the 1000 genome reference panel,^13^ Haplotype Reference Consortium, or the UK10K using IMPUTE4 software.^14^ Meta-analysis was performed using inverse-variance method and Cochran’s Q test (to estimate heterogeneity) with METAL.^15,16^ Age, sex, and the first twenty principal components were included as covariates in logistic regression models to identify IA-associated SNPs as previously described.^17^ Threshold for genome-wide significance was defined after Bonferroni correction for the total number of SNPs tested (p ≤ 5.0×10^−8^). Our cohort contained patients of both European and East-Asian ancestry. Assuming a disease prevalence for IA of 5%,^18^ rs73392700 allele frequency of 0.12 (the most statistically-significant SNP identified in our GWAS), and genotype relative risk of 1.5, our study is appropriately powered to 1.0 (GAS power calculator from the University of Michigan, csg.sph.umich.edu). STREGA reporting guidelines for genetic association studies were followed.^19^

### Functional genomics analysis

The latest data release of GTEx (version 8) was used.^20^ The GTEx portal (gtexportal.org) was used for data visualization. Expression quantitative trait loci (eQTL) analysis, which estimates the functional effect of a SNP/loci on neighboring gene expression, was performed as previously described.^11,20^ Splicing quantitative trait loci (sQTL) was used to determine the potential impact of a SNP on alternative splicing.^21^ In addition, we used Haploreg,^22^ a functional genomics tool to explore the function of variants in non-coding regions of the genome. Haploreg combines data from the 1000 genomes project, Roadmap Epigenomics, and ENCODE projects to estimate the effect of SNPs on functions such as chromatin state, protein-binding, and gene-expression. Data visualization was performed using gtexportal.org. Finally, we utilized single-cell RNA sequencing analysis of adult human brain endothelial and perivascular cells obtained from normal brain cortex biopsies obtained as previously described.^23^

## Results

Our cohort contained 84,353 individuals (7,843 rIA cases and 76,510 controls) across 24 published GWAS.^10^ Our GWAS identified 5 independent genetic loci reaching genome-wide significance for rIA (Figure 1A): rs73392700 (*CDKN2B*, p=4.84×10^−17^, OR=1.12), rs6841581 (*EDNRA*, p=3.52×10^−14^, OR=0.77), rs11661542 (*RBBP8*, p=3.18×10^−13^, OR=0.84), rs62516550 (*RP1*, p=2.94×10^−11^, OR=1.20), and rs12310399 (*FGD6*, p=3.19×10^−10^, OR=1.16). The *FGD6* loci, to our knowledge, has not been implicated in prior GWAS of rIA. We confirmed that population stratification and genomic control was adequately achieved (i.e., no early deviation of p-values from expected, lambda statistic 1.048) by generating a quantile-quantile plot (Figure 1B).

**Figure 1.**
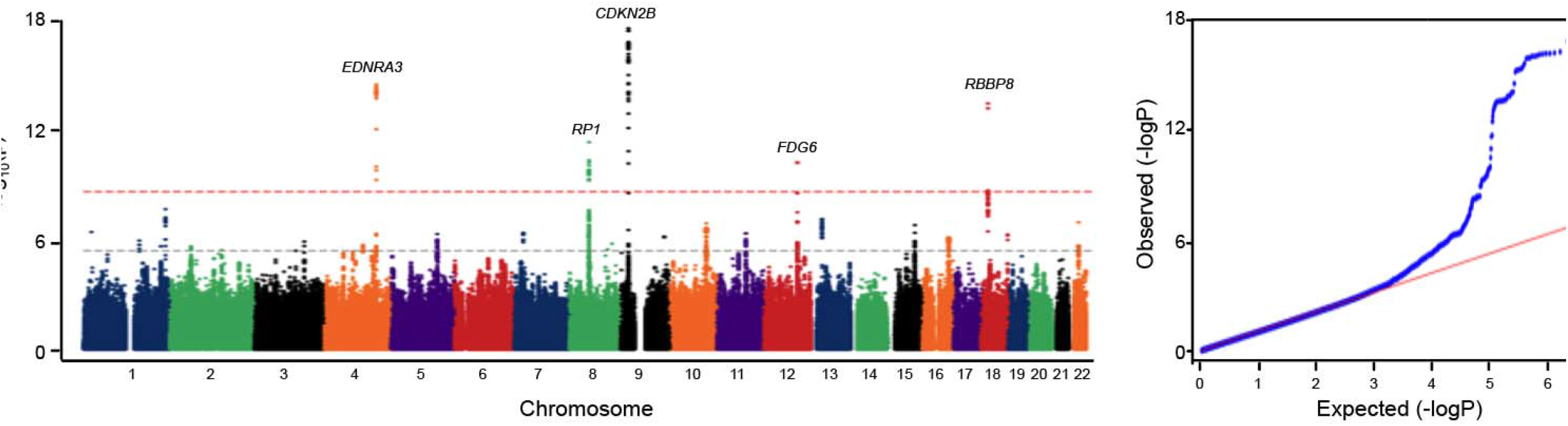
Genome-wide association study (GWAS) of ruptured intracranial aneurysms (rIA). (A) Manhattan plot of our rIA GWAS containing 84,353 individuals (7,843 rIA cases and 76,510 controls) and identify 5 independent genetic loci reaching genome-wide significance (p<5.0×10^−8^) for rIA: rs73392700 (*CDKN2B*, OR=1.12), rs6841581 (*EDNRA*, OR=0.77), rs11661542 (*RBBP8*, OR=0.84), rs62516550 (*RP1*, OR=1.20), and rs12310399 (*FGD6*, OR=1.16). The horizontal red line delineates the threshold for genome-wide significance (p = 5.0×10^−8^). (B) Quantile-Quantile (Q-Q) plot demonstrating adequate genomic control for population stratification (Lambda statistic 1.048). These data are visualized using the cd.hugemap.org.

Consistent with GWAS studies of complex and polygenic diseases, the vast majority of identified genetic risk factors are located in non-coding regions of the genome, making causal inference of disease mechanisms challenging.^24^ To overcome these challenges and delineate the functional consequences of rIA-associated single-nucleotide polymorphisms (SNPs), we analyzed the summative contribution of rIA associated SNPs across ∼20,000 individual genes using gene-burden analysis.^25^ Determining gene-based, rather than variant-based, associations with rIA will facilitate ease of biologic interpretation and design/interpretation of mechanistic studies. Across all rIA-associated SNPs and genes tested, 958 genes were associated with rIA (p< 0.05). However, after multiple-testing correction, only *FGD6* (harboring 21 rIA-associated SNPs) reached transcriptome-wide significance.

To visualize the *FGD6* locus for rIA-associated SNPs, we present a LocusZoom plot (Figure 2). These data demonstrate the extent to which multiple variants in this locus, alone and in concert through linkage disequilibrium (LD), contribute to rIA risk. To delineate the effect of rIA-associated SNPs on gene expression, we present eQTL mapping across 49 tissue types using the Genotype-Tissue Expression Project (GTEx) portal^26^ and demonstrate that the most significant rIA-associated SNP in the *FGD6* loci (rs12310399) causes decreased *FGD6* gene expression most-significantly in arterial tissue (Figure 3). In addition, rs12310399 overlaps splicing QTLs for vezatin (*VEZT*, p< 8.20×10^−9^, skeletal muscle, gtexportal.org), a transmembrane protein in the adherens junction that binds to myosin VIIA^27^ and NADH ubiquinone oxidoreductase subunit A12 (*NDUFA12*, p< 8.20×10^−9^, skeletal muscle, gtexportal.org), a gene previously implicated in IA biology.^28^ These data provide additional support for this locus and suggests that multiple mechanisms are likely contributing to rIA risk.

**Figure 2.**
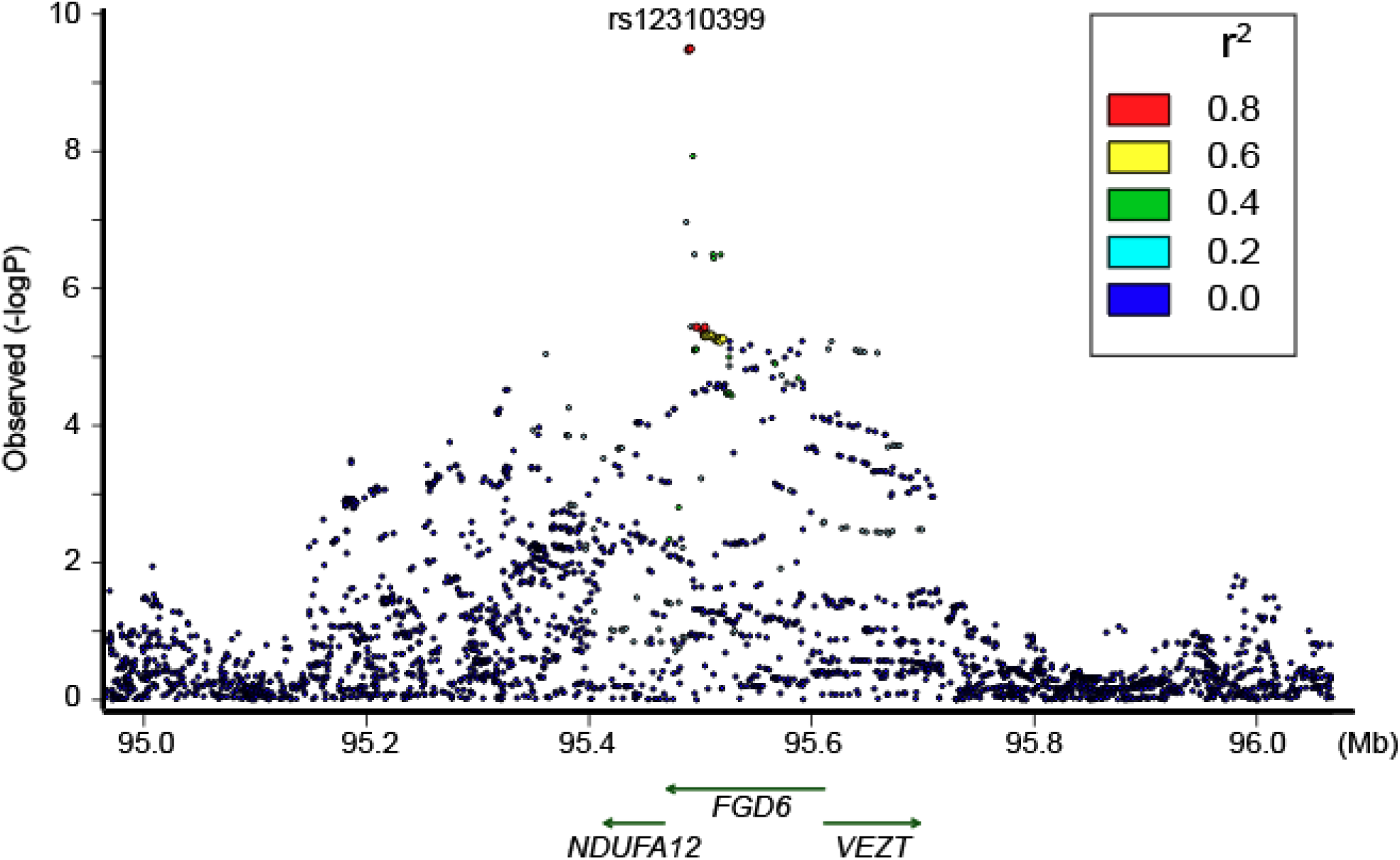
LocusZoom plot after fine-mapping of rIA-associated SNPs in the *FGD6* locus.

**Figure 3.**
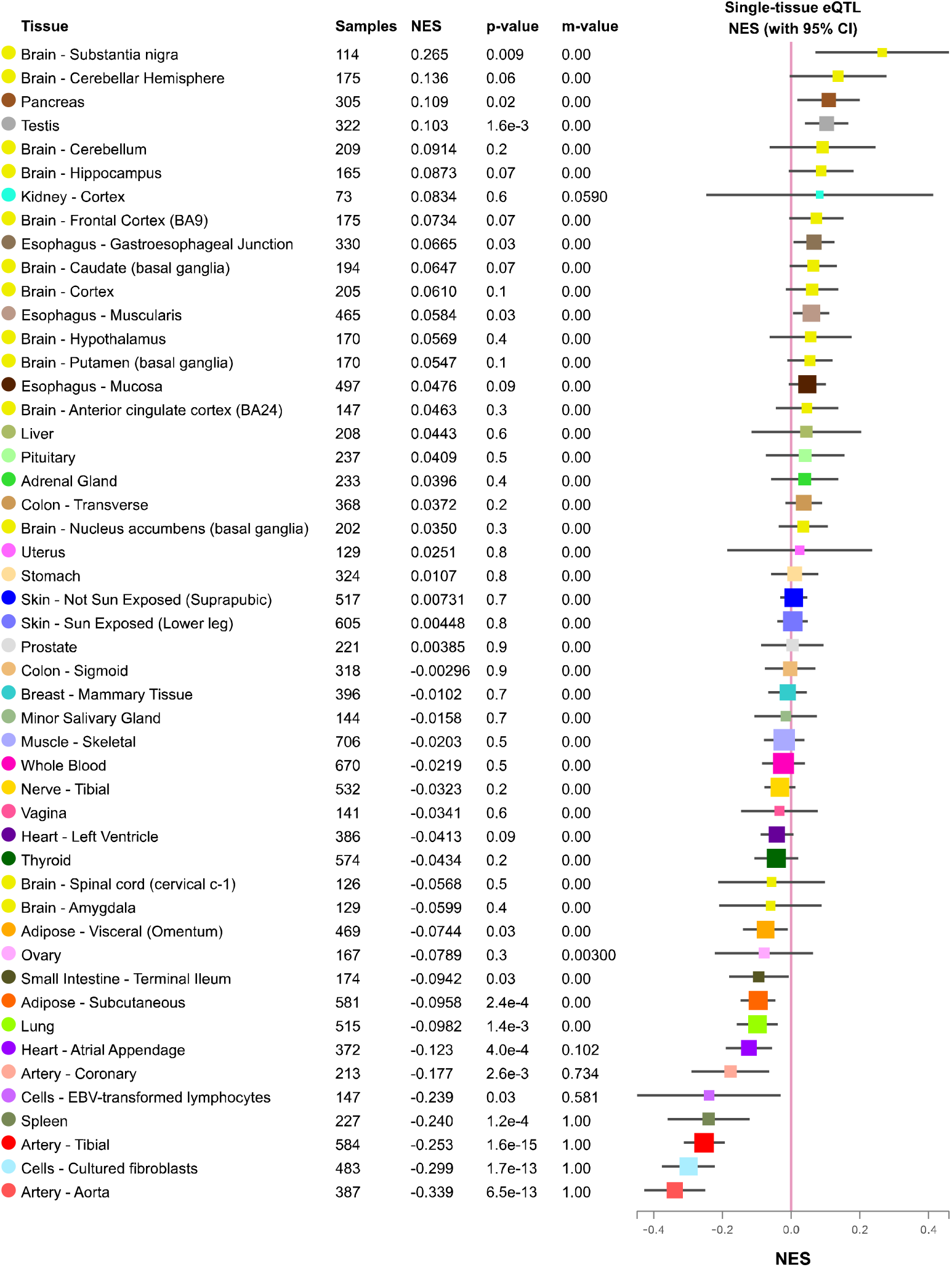
Expression quantitative trait loci (eQTL) analysis of the *FGD6* locus sentinel variant (rs12310399) across 49 tissues in GTEx. NES = normalized enrichment score. The m-value is the posterior probability from MetaSoft which is the probability than an eQTL effect exists in the tested cross-tissue analysis ^43,44^. A m-value < 0.01 is highly suggestive that an eQTL effect does not exist, whereas a m-value > 0.9 is highly predictive of an eQTL effect. Data were visualized using gtexportal.org.

While identification of new rIA-associated genes is important, it is the function of that gene in relevant tissues/cells that will expand our pathophysiologic understanding of IA formation and rupture. To further delineate the cellular origin and functional consequences of *FGD6* expression in the cerebrovascular system, we utilize scRNA-seq data derived from normal adult human brain endothelial and perivascular cells as previously described.^23^ We find that *FGD6* is highly expressed in 1 of 3 arterial lineages, venules, and veins but *FGD6* expression is largely undetectable across 12 perivascular cell lineages (Figure 5A-B). These findings will facilitate additional mechanistic studies aimed at understanding the molecular basis of *FGD6* dysfunction underlying rIA.

**Figure 4.**
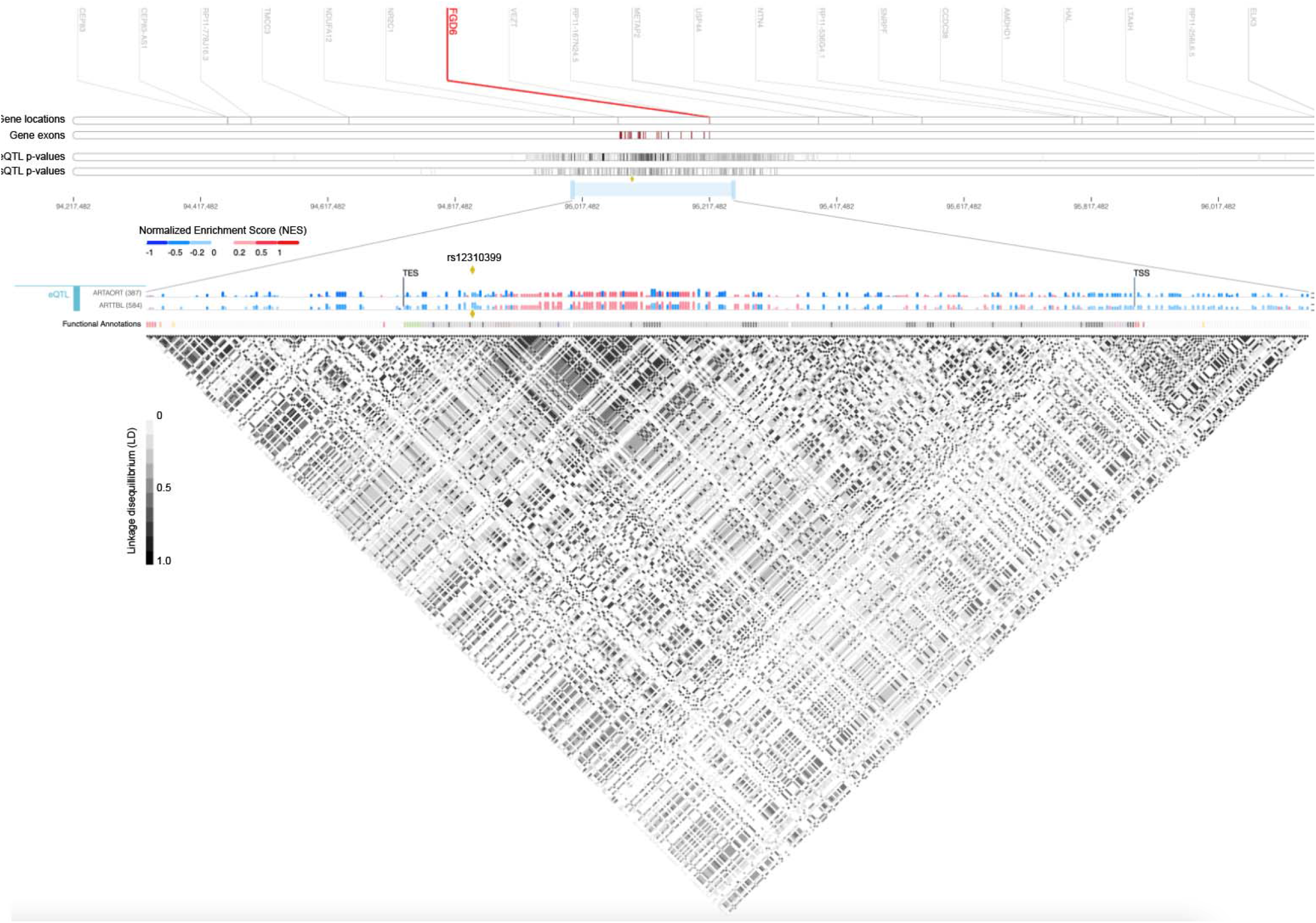
Functional mapping of the *FGD6* locus in arterial tissue. Expression quantitative trait loci (eQTL), splicing quantitative trait loci (sQTL), and linkage disequilibrium (LD) mapping of the FGD6 locus. Abbreviations are as follows: Transcription end site (TES), Transcription start site (TSS), Aortic artery (ARTORT), Tibial artery (ARTTBL). Data were visualized using gtexportal.org.

**Figure 5.**
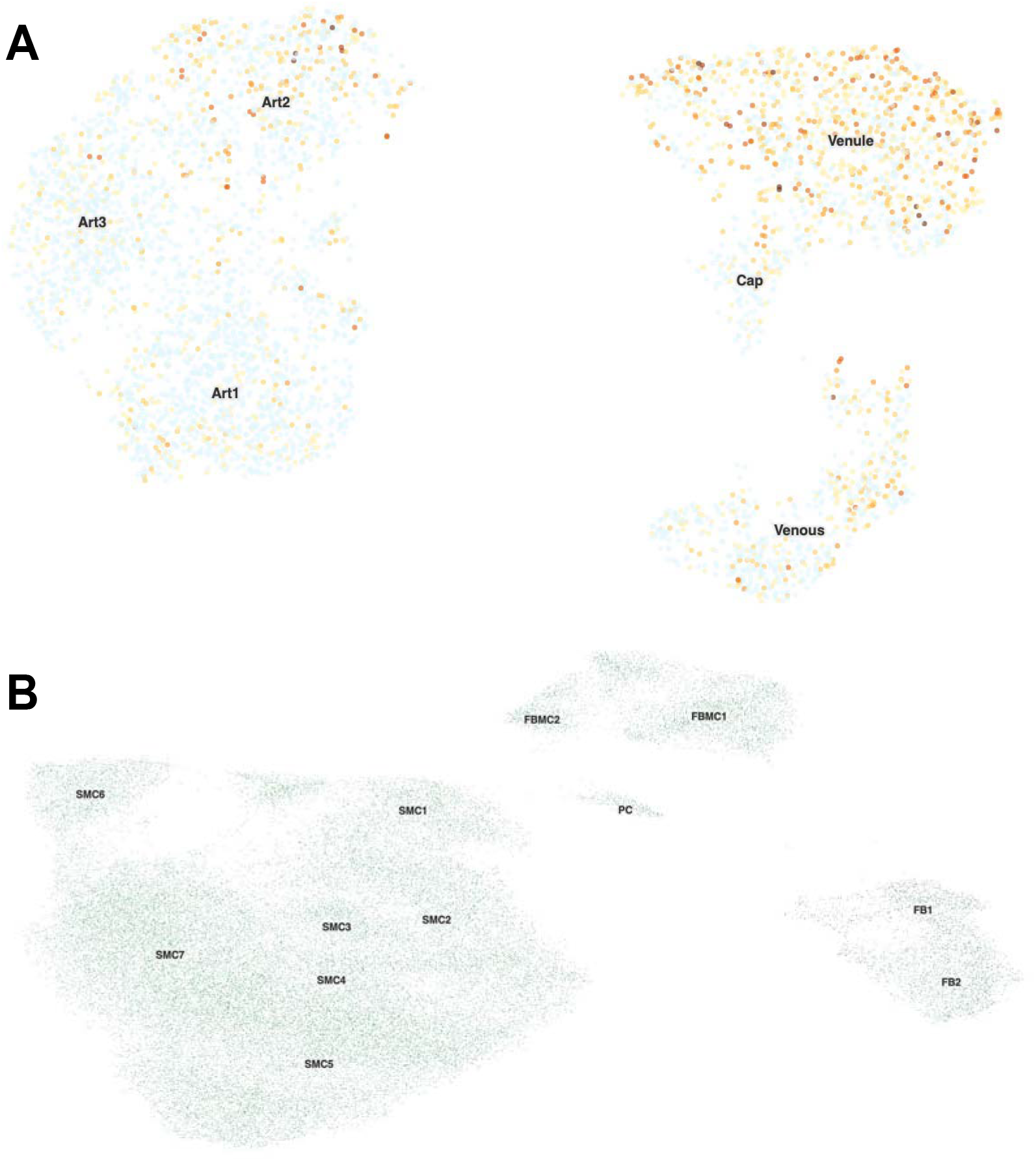
Single-cell RNA sequencing (scRNA-seq) analysis of *FGD6* across adult human brain (A) endothelial and (B) perivascular cells obtained from normal resected brain tissue. Endothelial cells were characterized into 3 arterial lineages (Art1-3), capillary (cap), venous, and venule cell-lineages. Blue dots indicate undetectable expression of *FGD6*, and increasingly dark shades of orange indicate higher expression of *FGD6*. Perivascular cells were partitioned into 7 smooth muscle (SMC1-7), 2 fibroblast (FB), 2 fibromyocyte (FbM1-2), and 1 pericyte (PC) lineage. Green dots indicate undetectable expression of *FGD6* and red dotes indicate higher expression of *FGD6*. UMAP data visualization and gene-expression markers for cell populations was performed using the UCSC Cell Browser.

## Discussion

We identify 5 independent genetic loci reaching genome-wide significance for rIA, including a novel locus containing *FGD6* not previously implicated in rIA biology. Gene-burden analysis provides convergent evidence supporting the *FGD6* gene in rIA risk. Functional characterization indicates that genetic variation in *FGD6* loci contributes to rIA risk by decreasing *FGD6* gene expression in arterial tissue, Utilization of single-cell RNA sequencing analysis of human brain endothelial and perivascular cells ^23^ characterizes *FGD6* expression across rIA-relevant neurovascular cell types where alterations in *FGD6* expression may alter cell identity and function. These data will inform detailed mechanistic analysis of *FGD6* in rIA biology and perhaps even treatment strategies. These data suggest that variable expressivity of *FDG6* causes a range of clinical features from Mendelian disease to sporadic rIA.

FGD6 (FYVE RhoGef and Ph domain containing protein 6) is a RhoGTPase that activates CDC42, a cell cycle-associated gene, and FGD6 has been shown to play a role in regulating angiogenesis and actin cytoskeleton rearrangement,^29^ functions consistent with rIA. Functional analysis indicates that *FGD6* regulates angiogenesis and actin cytoskeleton rearrangement.^29^ Thus, it is reasonable to posit that *FGD6*-dependent alterations in blood vessel integrity and/or formation may, at least in part, contribute to rIA. *FGD6* mediated genetic susceptibility coupled with clinical risk factors (hypertension, smoking, etc.) converge to contribute summative risk for rIA. Whole-gene deletion or introduction of FGD6 disease-causing mutations disrupts endothelial cell network formation which could be rescued by introduction of the wild-type gene product.

While the focus of this manuscript is on identification and characterization of the *FGD6* loci, multiple genetic loci are contributing to rIA (Figure 1). Although the magnitude of effect is relatively small for the lead SNP in the *FGD6* loci (OR= 1.16), the clinical consequences of genetic-testing underlying rIA are more likely to be representing through assessment of polygenic risk score (PRS).^30^ The weighted contribution of multiple loci through creation of a PRS will improve our understanding of rIA genetic architecture.^31^ In addition, combining PRS with important clinical and radiologic features of patients with rIA will improve translational of these genetic findings to clinical care. As our ability to acquire, analyze, and interpret genetic information improves, we surmise that translating genetic information into clinical practice will become streamlined.

Our cohort is the largest studies of rIA to date, but additional genetic data from diverse populations will increase power to detect additional risk loci and improve applicability of the genetic information across diverse populations. One limitation of array based GWAS is our relative inability to include rare variants, although detailed analysis of IA rare variants has been performed elsewhere.^32,33^ Multiple studies of rIA have implicated common variants underling IA risk.^10,28,34-39^ Thus, it is likely that both common and rare variants in both coding and non-coding regions confer risk to IA, alone and in concert. In addition, recent studies suggest that somatic mutations may play a role in IA pathology,^40,41^ which we did not assess herein, but may play a significant role in rIA pathogenesis. Analysis of lesional tissue acquired through advances in endoluminal biopsy will promote our understanding of somatic contribution to rIA biology.^42^ Integration of clinical and aneurysm-specific factors (size, location, etc.) will enable personalized medicine approaches to IA treatment and expand the clinician’s armamentarium.

Here we perform a large genetic study of rIA and identify and characterize a previously unknown risk locus for rIA containing *FGD6*. Since the genetic contribution to rIA risk does not change (excluding somatic mutations), approaches to quantify genetic risk of rIA may be extremely valuable for clinical treatment. Integration of clinical and radiologic IA factors with genetic data will further improve our ability to comprehensively evaluate rIA risk. Understanding the genetic determinants of rIA may inform gene-based therapies aimed at IA remodeling or obliteration. Strategies to incorporate genetic information into clinical care will inform personalized medicine approaches to manage IA.

## Data Availability

All data produced in the present study are available upon reasonable request to the authors.

## Acknowledgements

The authors would like to thank the creators of the Cerebrovascular Disease Knowledge Portal. In addition, we would like to thank GTEx which is supported by the Common Fund of the Office of the Director of the NIH, the National Cancer Institute (NCI), the National Human Genome Research Institute (NHGRI), the National Heart, Lung, and Blood Institute (NHLBI), the National Institute on Drug Abuse (NIDA), the National Institute of Mental Health (NIMH), and the National Institute of Neurological Disorders and Stroke (NINDS). We would also like to thank Haploreg (Broad Institute) - which also utilizes Roadmap Epigenomics, 1000 Genomes Project, and ENCODE data - for use of their resource.

## References

1. Lawton MT, Vates GE. Subarachnoid Hemorrhage. N Engl J Med. 2017;377(3):257–266.

2. Macdonald RL, Schweizer TA. Spontaneous subarachnoid haemorrhage. Lancet. 2017;389(10069):655–666.

3. Korja M, Kaprio J. Controversies in epidemiology of intracranial aneurysms and SAH. Nat Rev Neurol. 2016;12(1):50–55.

4. Schievink WI. Intracranial Aneurysms. New England Journal of Medicine. 1997;336(1):28–40.

5. Teunissen LL, Rinkel GJ, Algra A, van Gijn J. Risk factors for subarachnoid hemorrhage: a systematic review. Stroke. 1996;27(3):544–549.

6. Ilinca A, Samuelsson S, Piccinelli P, Soller M, Kristoffersson U, Lindgren AG. A stroke gene panel for whole-exome sequencing. European journal of human genetics : EJHG. 2018.

7. Zhou S, Dion PA, Rouleau GA. Genetics of Intracranial Aneurysms. Stroke. 2018;49(3):780–787.

8. Vlak MH, Algra A, Brandenburg R, Rinkel GJ. Prevalence of unruptured intracranial aneurysms, with emphasis on sex, age, comorbidity, country, and time period: a systematic review and meta-analysis. The Lancet Neurology. 2011;10(7):626–636.

9. Kleinloog R, van ‘t Hof FN, Wolters FJ, et al. The association between genetic risk factors and the size of intracranial aneurysms at time of rupture. Neurosurgery. 2013;73(4):705–708.

10. Bakker MK, van der Spek RAA, van Rheenen W, et al. Genome-wide association study of intracranial aneurysms identifies 17 risk loci and genetic overlap with clinical risk factors. Nat Genet. 2020;52(12):1303–1313.

11. Hale AT, Akinnusotu O, He J, et al. Genome-Wide Association Study Identifies Genetic Risk Factors for Spastic Cerebral Palsy. Neurosurgery. 2021;89(3):435–442.

12. Hale AT, Bastarache L, Morales DM, Wellons JC, 3rd, Limbrick DD, Jr., Gamazon ER. Multi-omic analysis elucidates the genetic basis of hydrocephalus. Cell Rep. 2021;35(5):109085.

13. Fairley S, Lowy-Gallego E, Perry E, Flicek P. The International Genome Sample Resource (IGSR) collection of open human genomic variation resources. Nucleic Acids Res. 2020;48(D1):D941–d947.

14. Bycroft C, Freeman C, Petkova D, et al. The UK Biobank resource with deep phenotyping and genomic data. Nature. 2018;562(7726):203–209.

15. Willer CJ, Li Y, Abecasis GR. METAL: fast and efficient meta-analysis of genomewide association scans. Bioinformatics. 2010;26(17):2190–2191.

16. Hale AT, He J, Akinnusotu O, et al. Genome-wide association study reveals genetic risk factors for trigeminal neuralgia. medRxiv. 2021:2021.2002.2008.21251349.

17. Hale AT, Zhou D, Sale RL, et al. The genetic architecture of human infectious diseases and pathogen-induced cellular phenotypes. medRxiv. 2021:2020.2007.2019.20157404.

18. Caranci F, Briganti F, Cirillo L, Leonardi M, Muto M. Epidemiology and genetics of intracranial aneurysms. Eur J Radiol. 2013;82(10):1598–1605.

19. Little J, Higgins JP, Ioannidis JP, et al. STrengthening the REporting of Genetic Association Studies (STREGA): an extension of the STROBE statement. PLoS medicine. 2009;6(2):e22.

20. The GTEx Consortium atlas of genetic regulatory effects across human tissues. Science (New York, NY). 2020;369(6509):1318–1330.

21. Li YI, Knowles DA, Humphrey J, et al. Annotation-free quantification of RNA splicing using LeafCutter. Nature genetics. 2018;50(1):151–158.

22. Ward LD, Kellis M. HaploReg: a resource for exploring chromatin states, conservation, and regulatory motif alterations within sets of genetically linked variants. Nucleic Acids Res. 2012;40(Database issue):D930–934.

23. Winkler EA, Kim CN, Ross JM, et al. A single-cell atlas of the normal and malformed human brain vasculature. Science (New York, NY). 2022:eabi7377.

24. Tak YG, Farnham PJ. Making sense of GWAS: using epigenomics and genome engineering to understand the functional relevance of SNPs in non-coding regions of the human genome. Epigenetics Chromatin. 2015;8:57.

25. de Leeuw CA, Mooij JM, Heskes T, Posthuma D. MAGMA: generalized gene-set analysis of GWAS data. PLoS Comput Biol. 2015;11(4):e1004219.

26. Human genomics. The Genotype-Tissue Expression (GTEx) pilot analysis: multitissue gene regulation in humans. Science (New York, NY). 2015;348(6235):648–660.

27. Guo X, Jing C, Li L, et al. Down-regulation of VEZT gene expression in human gastric cancer involves promoter methylation and miR-43c. Biochemical and biophysical research communications. 2011;404(2):622–627.

28. Yasuno K, Bakircioglu M, Low SK, et al. Common variant near the endothelin receptor type A (EDNRA) gene is associated with intracranial aneurysm risk. Proc Natl Acad Sci U S A. 2011;108(49):19707–19712.

29. Huang L, Zhang H, Cheng CY, et al. A missense variant in FGD6 confers increased risk of polypoidal choroidal vasculopathy. Nat Genet. 2016;48(6):640–647.

30. GWAS to the people. Nat Med. 2018;24(10):1483.

31. Dudbridge F. Power and predictive accuracy of polygenic risk scores. PLoS Genet. 2013;9(3):e1003348.

32. Kurki MI, Gaál EI, Kettunen J, et al. High risk population isolate reveals low frequency variants predisposing to intracranial aneurysms. PLoS Genet. 2014;10(1):e1004134.

33. Barak T, Ristori E, Ercan-Sencicek AG, et al. PPIL4 is essential for brain angiogenesis and implicated in intracranial aneurysms in humans. Nat Med. 2021;27(12):2165–2175.

34. Bilguvar K, Yasuno K, Niemelä M, et al. Susceptibility loci for intracranial aneurysm in European and Japanese populations. Nat Genet. 2008;40(12):1472–1477.

35. Yasuno K, Bilguvar K, Bijlenga P, et al. Genome-wide association study of intracranial aneurysm identifies three new risk loci. Nat Genet. 2010;42(5):420–425.

36. Foroud T, Koller DL, Lai D, et al. Genome-wide association study of intracranial aneurysms confirms role of Anril and SOX17 in disease risk. Stroke. 2012;43(11):2846–2852.

37. Low SK, Takahashi A, Cha PC, et al. Genome-wide association study for intracranial aneurysm in the Japanese population identifies three candidate susceptible loci and a functional genetic variant at EDNRA. Human molecular genetics. 2012;21(9):2102–2110.

38. Foroud T, Lai D, Koller D, et al. Genome-wide association study of intracranial aneurysm identifies a new association on chromosome 7. Stroke. 2014;45(11):3194–3199.

39. Laarman MD, Vermunt MW, Kleinloog R, et al. Intracranial Aneurysm-Associated Single-Nucleotide Polymorphisms Alter Regulatory DNA in the Human Circle of Willis. Stroke. 2018;49(2):447–453.

40. Chenbhanich J, Hu Y, Hetts S, et al. Segmental overgrowth and aneurysms due to mosaic PDGFRB p.(Tyr562Cys). Am J Med Genet A. 2021;185(5):1430–1436.

41. Karasozen Y, Osbun JW, Parada CA, et al. Somatic PDGFRB Activating Variants in Fusiform Cerebral Aneurysms. Am J Hum Genet. 2019;104(5):968–976.

42. Winkler EA, Wu D, Gil E, et al. Endoluminal Biopsy for Molecular Profiling of Human Brain Vascular Malformations. Neurology. 2022.

43. Han B, Eskin E. Random-effects model aimed at discovering associations in meta-analysis of genome-wide association studies. Am J Hum Genet. 2011;88(5):586–598.

44. Han B, Eskin E. Interpreting meta-analyses of genome-wide association studies. PLoS Genet. 2012;8(3):e1002555.

